# Association between vaccination status and reported incidence of post-acute COVID-19 symptoms in Israel: a cross-sectional study of patients tested between March 2020 and November 2021

**DOI:** 10.1101/2022.01.05.22268800

**Authors:** Paul Kuodi, Yanay Gorelik, Hiba Zayyad, Ofir Wertheim, Karine Beiruti Wiegler, Kamal Abu Jabal, Amiel A. Dror, Saleh Nazzal, Daniel Glikman, Michael Edelstein

## Abstract

**Background:** Long COVID is a post-severe acute respiratory syndrome coronavirus 2 (SARS-CoV-2) infection syndrome characterised by not recovering for several weeks or months following the acute episode. The effectiveness of COVID-19 vaccines against long-term symptoms of COVID-19 is not well understood. We determined whether vaccination was associated with the incidence of reporting long-term symptoms post-SARS-CoV-2 infection

**Methods:** We invited individuals who were PCR tested for SARS-CoV-2 infection at participating hospitals between March 2020-November 2021 to fill an online questionnaire that included baseline demographics, details of their acute episode and information about symptoms they were currently experiencing. Using binomial regression, we compared vaccinated individuals with those unvaccinated and those uninfected in terms of self-reported symptoms post-acute infection.

**Results:** We included 951 infected and 2437 uninfected individuals. Of the infected, 637(67%) were vaccinated. The most commonly reported symptoms were; fatigue (22%), headache (20%), weakness (13%), and persistent muscle pain (10%). After adjusting for follow-up time and baseline symptoms, those who received two doses less likely than unvaccinated individuals to report any of these symptoms by 64%, 54%, 57%, and 68% respectively, (Risk ratios 0.36, 0.46, 0.43, 0.32, p<0.04 in the listed sequence). Those who received two doses were no more likely to report any of these symptoms than individuals reporting no previous SARS-CoV-2 infection.

**Conclusions:** Vaccination with at least two doses of COVID-19 vaccine was associated with a substantial decrease in reporting the most common post-acute COVID-19 symptoms, bringing it back to baseline. Our results suggest that, in addition to reducing the risk of acute illness, COVID-19 vaccination may have a protective effect against long COVID.

## Introduction

Long coronavirus disease 2019 (Long COVID), also known as post-COVID-19 syndrome, is an emerging and complex health problem that remains poorly characterised. In October 2021, the World Health Organization (WHO) defined long COVID as “A condition which occurs in individuals with a history of probable or confirmed severe acute respiratory syndrome coronavirus 2 (SARS-CoV-2) infection, usually three months from the onset of COVID-19 with symptoms that last for at least two months and cannot be explained by an alternative diagnosis. Common symptoms include fatigue, shortness of breath, and cognitive dysfunction but also others involving the musculo-skeletal, cardiac and central nervous systems, which generally have an impact on everyday functioning” [1]. Symptoms of long COVID may fluctuate or relapse over time. Patients purported to have long COVID have reported a wide range of symptoms but the most prevalent symptoms include; fatigue (approximately 58%), shortness of breath (24%), joint pain (19%), chest pain (16%) [2, 3], headache (44%), palpitations (11%), physical limitations, depression (12%) and insomnia (11%) [3, 4]. These symptoms may emerge after the initial recovery from an acute COVID-19 episode, or be persistent symptoms that do not resolve following the initial COVID-19 illness [5].

Vaccination against SARS-CoV-2 infection is one of the most important interventions deployed to mitigate the COVID-19 pandemic. As of October 2021, the WHO has listed the Pfizer/BioNTech, AstraZeneca-SK Bio, Sinopharm, Serum Institute of India, Janssen, and Moderna vaccines for emergency use under the Emergency Use Authorisation (EUA) [6]. In August 2021, the Food and Drugs Administration (FDA) approved the Pfizer/BioNTech vaccine as the first vaccine for use to prevent COVID-19 in individuals sixteen years and older [7].

The first COVID-19 vaccine campaign with a WHO-approved vaccine started in December 2020, following the authorisation of the Pfizer/BioNTech mRNA vaccine by the FDA for emergency use. By January 2022, over 58% of the world population had received at least one dose of the EUA COVID-19 vaccines, accounting for 9.2 billion COVID-19 doses [8]. Available evidence demonstrates that COVID-19 vaccines are effective in preventing severe complications of COVID-19 and death [9], including in cases of infection with the Delta and Omicron variants of SARS-CoV-2 [10-12], variants that are more contagious than the ancestral SARS-CoV-2 variants [12,13]. In Israel, the COVID-19 vaccination campaign started in December 2020, and by January 2022, approximately 63.6% of the entire population had received a at least two doses, mainly with the BNT162b2 mRNA vaccine [14] and 45.6% had received a third dose, available in Israel from June 2021. The third dose demonstrated high effectiveness against severe outcomes in the context of waning immunity six months post-priming course [14]. During the study period, individuals previously infected with SARS-CoV-2 were eligible for a single dose of the BNT162b2 mRNA vaccine only. All other individuals with no history of SARS-CoV-2 infection were eligible for a first and a second dose with eligibility for the second dose at three weeks from the date of the first dose.

Risk factors for developing long COVID have not been fully explored. So far, increasing age, pre-existing health conditions such as hypertension, obesity, psychiatric disorders, and immunosuppression have been associated with an increased risk of long COVID [15]. As the process to establish a more specific case definition of long COVID continues, little is known about the impact of COVID-19 vaccination on long COVID. A large prospective study reported an association between vaccination and lower self-reporting of symptoms beyond 28 days among SARS-CoV-2 infected individuals without reporting details on specific symptoms or duration [16]. Findings from one study (in preprint) suggests vaccination reduces the reporting of some, but not all post-acute COVID-19 sequelae 6 months post-infection [17]. Another study, albeit with a small sample and without a control group, reported a high proportion of COVID-19 breakthrough infections among patients with long COVID symptoms [18]. An additional study reported that 20% of individuals infected post-vaccination developed post-COVID-19 symptoms beyond 6 weeks, however the investigators did not carry out baseline comparisons [19]. In the current study, we aimed to determine whether vaccination status was associated with reporting specific long-term symptoms post-COVID-19 infection.

## Methods

### Study design and participants

The current study reports results of a cross-sectional study nested in a prospective longitudinal cohort study to assess risk of and risk factors for long-term physical, mental, and psychosocial consequences of COVID-19. All individuals over the age of 18 who were tested for SARS-CoV-2 infection by reverse transcription polymerase chain reaction (RT-PCR) between 15^th^ March 2020 and 15^th^ November 2021 in the three major government hospitals in Northern Israel, namely Ziv Medical Centre, Padeh-Poriya Medical Centre, and Galilee Medical Centre, were eligible to join the study regardless of the test result. Using available patient telephone records, individuals were invited to participate in the study between July 16^th^ and November 18^th^ 2021, through a Short Message Service (SMS) containing an invitation with a link to an online survey available in four commonly spoken languages in Israel: Hebrew, Arabic, Russian, and English. Two reminders to complete the survey were sent to non-responders. In this study, we first restricted the analysis to individuals who reported being infected with SARS-CoV-2 and diagnosed by PCR in order to compare vaccinated to unvaccinated individuals in terms of reported long-term outcomes. Because the most commonly reported long-term outcomes reported by COVID-19 patients are not specific to COVID-19, we then compared the frequency of reported symptoms among infected and vaccinated individuals to never infected individuals.

### Measurement tools

The data collection tool was modified from the International Severe Acute Respiratory and emerging Infection Consortium (ISARIC) COVID-19 tools developed by a global follow-up working group and informed by a wide range of global stakeholders with expertise in clinical research, outbreak research, infectious disease, epidemiology, respiratory, critical care, rehabilitation, neurology, psychology, rheumatology, cardiology, oncology, and public health medicine [20]. The ISARIC questionnaire was adapted to the Israeli context. Study participants were asked to select from a list all symptoms they were currently experiencing.

### Data collected

The online survey included information about socio-demographic status (place of residence, age, sex, ethnicity/religious affiliation, education level and income), baseline health status: (Body mass index [BMI] and chronic conditions), information related to the clinical experience and symptoms experienced during the initial COVID-19 diagnosis, SARS-CoV-2 test results, vaccination status, number of doses, type of vaccine and date of administration. Participants were also asked to report symptoms experienced at the time of filling the questionnaire.

### Outcomes

The primary outcome in this study was the proportion, overall, and in specific age groups, of participants reporting selected health outcomes according to vaccination and infection status. It was not possible to determine vaccination status at the time of infection. As a result, some individuals were infected prior to vaccination while others were vaccinated after. Because of vaccination policy in Israel at the time that recommended a single dose for previously infected individuals, it is likely that most individuals who received a single dose were infected prior to vaccination whereas those who received two doses were infected after receiving their vaccines. The complete list of symptoms can be found in Table 1.

**Table 1.**
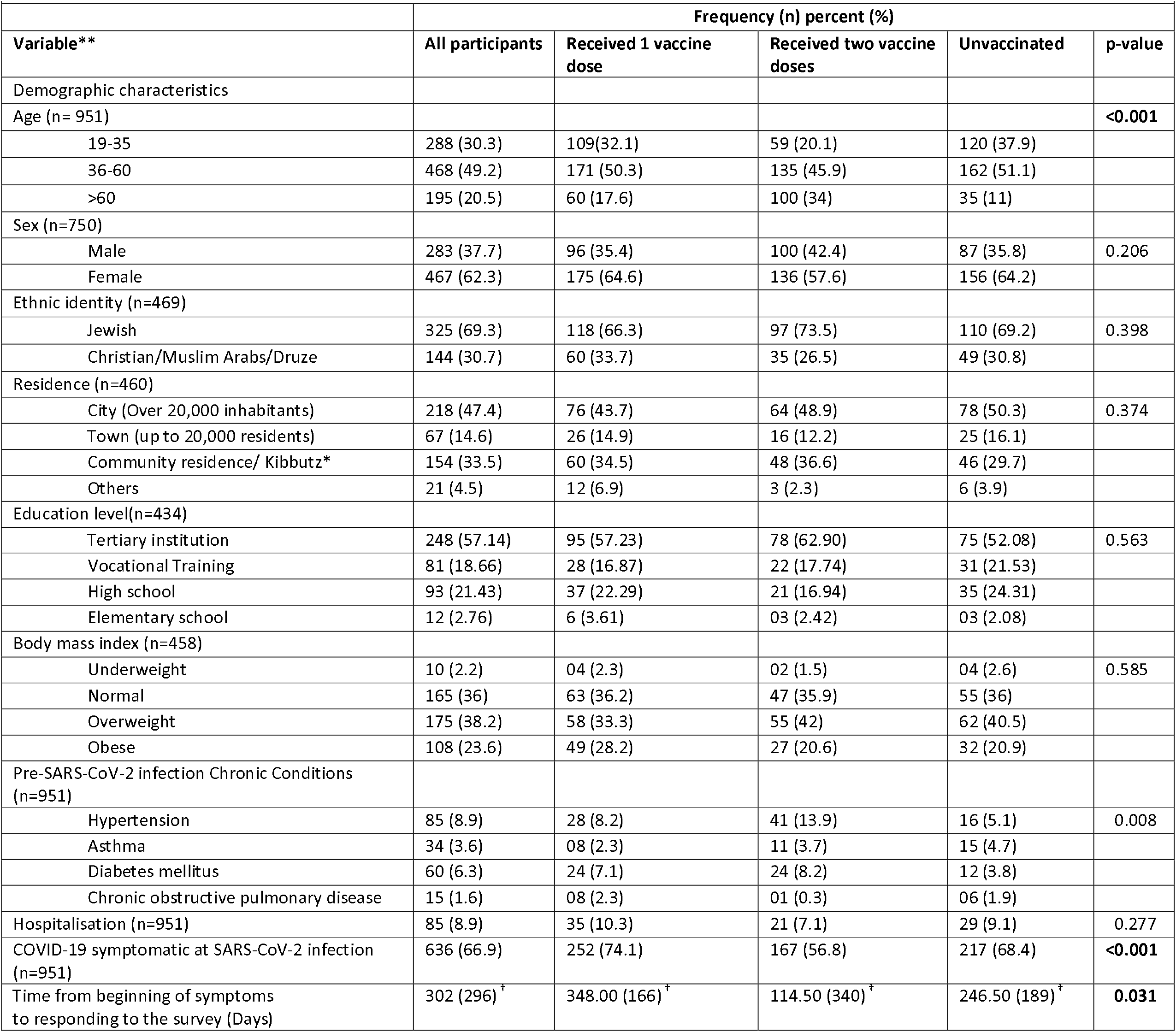
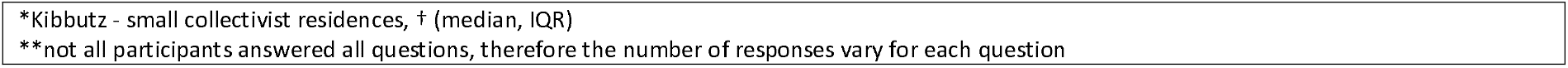
Baseline characteristics of the study population.

| Table 1. Baseline characteristics of the study population |  |  |  |  |  |
| --- | --- | --- | --- | --- | --- |
|  | Frequency (n) percent (%) |  |  |  |  |
| Variable** | All participants | Received 1 vaccine dose | Received two vaccine doses | Unvaccinated | p-value |
| Demographic characteristics |  |  |  |  |  |
| Age (n= 951) |  |  |  |  | <0.001 |
| 19-35 | 288 (30.3) | 109(32.1) | 59 (20.1) | 120 (37.9) |  |
| 36-60 | 468 (49.2) | 171 (50.3) | 135 (45.9) | 162 (51.1) |  |
| >60 | 195 (20.5) | 60 (17.6) | 100 (34) | 35 (11) |  |
| Sex (n=750) |  |  |  |  |  |
| Male | 283 (37.7) | 96 (35.4) | 100 (42.4) | 87 (35.8) | 0.206 |
| Female | 467 (62.3) | 175 (64.6) | 136 (57.6) | 156 (64.2) |  |
| Ethnic identity (n=469) |  |  |  |  |  |
| Jewish | 325 (69.3) | 118 (66.3) | 97 (73.5) | 110 (69.2) | 0.398 |
| Christian/Muslim Arabs/Druze | 144 (30.7) | 60 (33.7) | 35 (26.5) | 49 (30.8) |  |
| Residence (n=460) |  |  |  |  |  |
| City (Over 20,000 inhabitants) | 218 (47.4) | 76 (43.7) | 64 (48.9) | 78 (50.3) | 0.374 |
| Town (up to 20,000 residents) | 67 (14.6) | 26 (14.9) | 16 (12.2) | 25 (16.1) |  |
| Community residence/ Kibbutz* | 154 (33.5) | 60 (34.5) | 48 (36.6) | 46 (29.7) |  |
| Others | 21 (4.5) | 12 (6.9) | 3 (2.3) | 6 (3.9) |  |
| Education level(n=434) |  |  |  |  |  |
| Tertiary institution | 248 (57.14) | 95 (57.23) | 78 (62.90) | 75 (52.08) | 0.563 |
| Vocational Training | 81 (18.66) | 28 (16.87) | 22 (17.74) | 31 (21.53) |  |
| High school | 93 (21.43) | 37 (22.29) | 21 (16.94) | 35 (24.31) |  |
| Elementary school | 12 (2.76) | 6 (3.61) | 03 (2.42) | 03 (2.08) |  |
| Body mass index (n=458) |  |  |  |  |  |
| Underweight | 10 (2.2) | 04 (2.3) | 02 (1.5) | 04 (2.6) | 0.585 |
| Normal | 165 (36) | 63 (36.2) | 47 (35.9) | 55 (36) |  |
| Overweight | 175 (38.2) | 58 (33.3) | 55 (42) | 62 (40.5) |  |
| Obese | 108 (23.6) | 49 (28.2) | 27 (20.6) | 32 (20.9) |  |
| Pre-SARS-CoV-2 infection Chronic Conditions (n=951) |  |  |  |  |  |
| Hypertension | 85 (8.9) | 28 (8.2) | 41 (13.9) | 16 (5.1) | 0.008 |
| Asthma | 34 (3.6) | 08 (2.3) | 11 (3.7) | 15 (4.7) |  |
| Diabetes mellitus | 60 (6.3) | 24 (7.1) | 24 (8.2) | 12 (3.8) |  |
| Chronic obstructive pulmonary disease | 15 (1.6) | 08 (2.3) | 01 (0.3) | 06 (1.9) |  |
| Hospitalisation (n=951) | 85 (8.9) | 35 (10.3) | 21 (7.1) | 29 (9.1) | 0.277 |
| COVID-19 symptomatic at SARS-CoV-2 infection (n=951) | 636 (66.9) | 252 (74.1) | 167 (56.8) | 217 (68.4) | <0.001 |
| Time from beginning of symptoms to responding to the survey (Days) | 302 (296)<sup>†</sup> | 348.00 (166)<sup>†</sup> | 114.50 (340)<sup>†</sup> | 246.50 (189)<sup>†</sup> | 0.031 |

\*Kibbutz - small collectivist residences, † (median, IQR)
\*\*not all participants answered all questions, therefore the number of responses vary for each question

### Statistical analysis

We compared the vaccinated and unvaccinated groups’ socio-demographic and health characteristics using Chi-square tests (for proportions) and Student’s t-tests (for means). We also compared the proportion of participants presenting with symptomatic disease at baseline using Chi-square tests. Proportions of long-term symptoms and selected health outcomes were calculated for each group with the total number of participants in each group taken as the denominator. We also compared the time elapsed between the onset of COVID-19 symptoms for symptomatic patients and the date of response to the survey in the two groups (vaccinated and unvaccinated) to establish whether they had comparable durations of follow-up.

A first series of binomial regression models were fitted to the data for the ten most commonly reported post-COVID-19 symptoms according to vaccination status. We adjusted for the difference in follow-up time and proportion of asymptomatic patients at the time of diagnosis between the groups. We then compared vaccinated and infected individuals to never infected individuals in terms of reported symptoms, also using binomial regression models. We used two distinct steps since it was not possible to adjust for follow-up time and symptoms at baseline among uninfected patients.

To take the anticipated age differences into account, the analysis was age-stratified and differences in the length of time from the beginning of symptoms to responding to the survey were adjusted for in the model. Vaccination status was recorded as either one dose or two doses. At the time of data collection, very few individuals had received a third dose and those who did were recorded as two doses.

Datasets were processed using Microsoft Excel and analysed using STATA version 15.

### Ethical considerations

The study received ethical approvals from the Ziv Medical Centre, Padeh-Poriya Medical Centre, and Galilee Medical Centre ethical committees, reference numbers; 0007-21-ZIV, 009-21-POR, and 0018-21-NHR, respectively.

## Results

### Characteristics of the study population

Of 79482 invitations to participate in the study, 3562 (4.5%) individuals over the age of 18 agreed to participate: 2437 who reported no previous SARS-CoV-2 infection and 1125 who did. We excluded 174 infected individuals from the study because they did not report their vaccination status.

Of the 951 infected individuals included in the study, 340 (36%) reported receiving a single dose and 294 (31%) reported having received at least two doses. Of the 951 infected individuals, 636 (67%) reported at least one symptom during COVID-19 diagnosis. Among the unvaccinated, 69% reported at least one symptom at COVID-19 diagnosis, vs. 57% among those who received two doses and 74% among those who received one dose only, (p<0.05, Table 1).

Compared with individuals vaccinated with one and two doses, unvaccinated participants were younger (52 and 44 vs. 39 years, respectively, p<0.001, Table 1), reflecting the COVID-19 vaccination patterns in the general population. Consequently, pre-existing chronic conditions were more frequently reported in the vaccinated group (p<0.05). The vaccinated and unvaccinated groups were comparable in terms of gender distribution and socio-economic characteristics (Table 1). The median time between acute illness and reporting symptoms was longer in the unvaccinated group compared to those who received two doses (8 vs. 4 months, p<0.05), likely reflecting the fact that individuals who received one dose were by and large infected before being vaccinated. Out of 951 infected participants, 85 (9%) reported having been hospitalized with comparable proportions in all groups (p=0.27, Table 1).

### Post-COVID-19 symptoms among vaccinated and unvaccinated participants

Of the 951 infected participants, 337 (35%) reported not fully recovering from the initial COVID-19 symptoms at follow-up. The most commonly reported symptoms at the time of follow-up were fatigue (22%), headache (20%), and weakness in arms or legs (13%) and persistent muscle pain (10%) (Table 2).

**Table 2.** Post COVID-19 symptoms among vaccinated and unvaccinated participants.

|  |  |  |  |  |
| --- | --- | --- | --- | --- |
|  | Number and proportion experiencing post COVID symptoms, (n (%)) |  |  |  |
| **Post COVID symptoms** | **All participants (n=951)** | **Received 1 vaccine dose (n=340)** | **Received two vaccine doses (n=294)** | **Unvaccinated (n=317)** |
| Fatigue | 208 (21.87) | 93 (27.35) | 33 (11.22) | 82 (25.87) |
| Headache | 190 (19.98) | 80 (23.53) | 41 (13.95) | 69 (21.77) |
| Weakness in arms or legs | 128 (13.46) | 57 (16.76) | 20 (6.08) | 51 (16.09) |
| Persistent muscle pain | 98 (10.30) | 45 (13.24) | 17 (5.78) | 36 (11.36) |
| Loss of concentration | 90 (9.46) | 44 (12.94) | 13 (4.42) | 33 (10.41) |
| Hair loss | 88 (9.25) | 43 (12.65) | 09 (3.06) | 36 (11.36) |
| Problem sleeping | 85 (8.94) | 42 (12.35) | 14 (4.76) | 29 (9.15) |
| Dizziness | 74 (7.78) | 30 (8.82) | 12 (4.08) | 32 (10.09) |
| Persistent cough | 70 (7.36) | 26 (7.65) | 20 (6.80) | 24 (7.57) |
| Shortness of breath | 68 (7.15) | 29 (8.53) | 14 (4.76) | 25 (7.89) |
| Loss of taste | 63 (6.62) | 20 (5.88) | 15 (5.10) | 28 (8.83) |
| Chest pains | 61 (6.41) | 24 (7.06) | 14 (4.76) | 23 (7.26) |
| Pins and needles | 60 (6.31) | 31 (9.12) | 06 (2.04) | 23 (7.26) |
| Palpitations | 57 (5.99) | 26 (7.65) | 12 (4.08) | 19 (5.99) |
| Depression and anxiety | 55 (5.78) | 19 (5.59) | 17 (5.78) | 19 (5.99) |
| Abdominal pain | 54 (5.68) | 24 (7.06) | 08 (2.72) | 22 (6.94) |
| Problems with balance | 52 (5.47) | 24 (7.06) | 07 (2.38) | 21 (6.62) |
| Inability to control body movement | 49 (5.15) | 22 (6.47) | 09 (3.06) | 18 (5.68) |
| Joint pain or swelling | 47 (4.94) | 22 (6.47) | 05 (1.70) | 20 (6.31) |
| Loss of smell | 41 (4.31) | 09 (2.65) | 09 (3.06) | 23 (7.26) |
| Loss of appetite | 40 (4.21) | 11 (3.24) | 12 (4.08) | 17 (5.36) |
| Pain on breathing | 40 (4.21) | 16 (4.71) | 08 (2.72) | 16 (5.05) |
| Nausea and vomiting | 37 (3.89) | 11 (3.24) | 07 (2.38) | 19 (5.99) |
| Constipation | 34 (3.58) | 12 (3.53) | 08 (2.72) | 14 (4.42) |
| Erectile dysfunction | 32 (3.36) | 12 (3.53) | 08 (2.72) | 12 (3.79) |
| Diarrhoea | 31 (3.26) | 11 (3.24) | 05 (1.70) | 15 (4.73) |
| Double vision | 29 (3.05) | 17 (5.00) | 03 (1.02) | 09 (2.84) |
| Problems speaking or communicating | 28 (2.94) | 15 (4.41) | 03 (1.02) | 10 (3.15) |
| Weight loss | 28 (2.94) | 06 (1.76) | 08 (2.72) | 14 (4.42) |
| Tremor / shakiness | 27 (2.84) | 11 (3.24) | 08 (2.72) | 08 (2.52) |
| Problems passing urine | 16 (1.68) | 06 (1.76) | 02 (0.68) | 08 (2.52) |
| Loss of sensation, one side of the body | 15 (1.58) | 07 (2.06) | 02 (0.68) | 06 (1.89) |
| Skin lumps or rashes | 13 (1.37) | 05 (1.47) | 02 (0.68) | 06 (1.89) |
| Problems swallowing or chewing | 12 (1.26) | 05 (1.47) | 03 (1.02) | 04 (1.26) |
| Blood clots in veins | 12 (1.26) | 05 (1.47) | 03 (1.02) | 04 (1.26) |
| Kidney problems | 12 (1.26) | 05 (1.47) | 02 (0.68) | 05 (1.58) |
| Fainting / blackouts | 10 (1.05) | 04 (1.18) | 04 (1.36) | 02 (0.63) |
| Seizures / fits | 10 (1.05) | 04 (1.18) | 03 (1.02) | 03 (0.95) |
| Heart attack & stroke | 03 (0.32) | 02 (1.87) | 01 (1.27) | 00 (0.00) |

### Risk of reporting post-COVID-19 symptoms according to vaccination status

Compared with unvaccinated participants, those who received two doses were 36-73% less likely to report eight of the ten most commonly reported symptoms (p<0.04 for all, Table 3). After adjusting for duration of follow-up and presence of symptoms at baseline, a 54-82% reduction in reporting symptoms among those who received two doses for seven of the ten most commonly reported symptoms was detected (Table 3). Infected individuals who received two doses of COVID-19 vaccine were no more likely to report any of these symptoms than individuals who reported never having been infected with SARS-CoV-2 (Table 3).

**Table 3.**
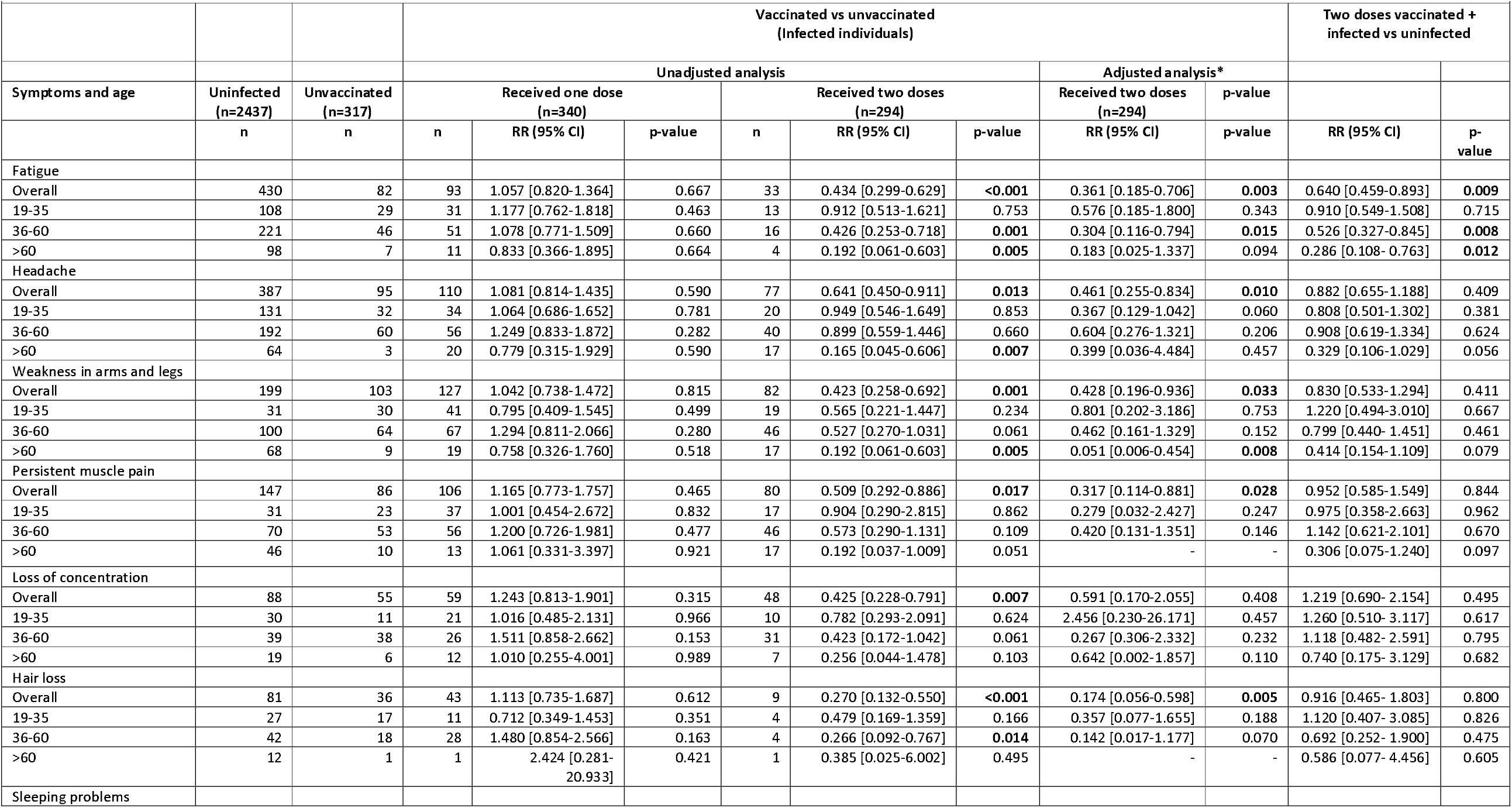
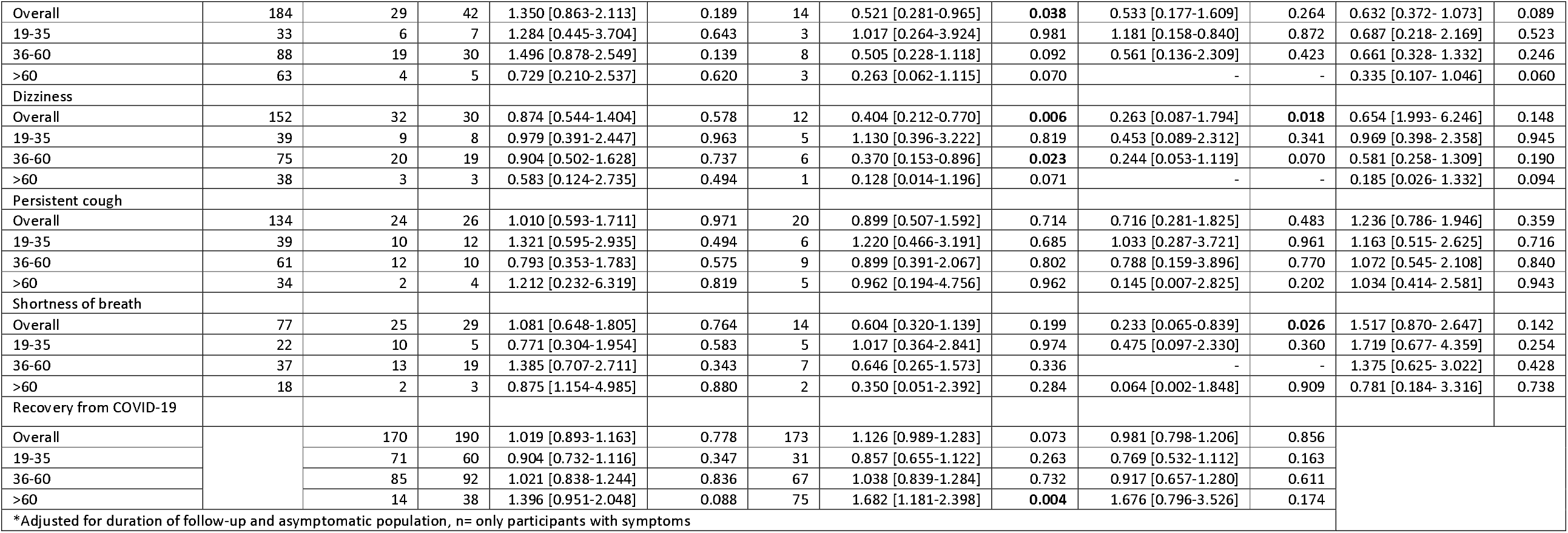
Crude and age-specific risk ratios of the most frequent post COVID symptoms among participants

In the unadjusted age-stratified analysis, differences in reported symptoms were primarily seen in the older age groups, in particular those aged above 60 years who were also 68% more likely to report feeling fully recovered compared with their unvaccinated peers of the same age (Table 3, p<0.004). Although the risk ratio remained high in the adjusted analysis (RR 1.7), the result was no longer significant, likely due to loss of statistical power.

## Discussion

Overall, receipt of two doses of COVID-19 vaccination was associated with a substantial decrease in reporting the most common post-COVID-19 symptoms. Individuals who had received two doses reported no more of these symptoms than never infected individuals. Our results therefore suggest that receiving two doses of COVID-19 vaccine, at least the BnT162b2 mRNA vaccine used in Israel brings the incidence of such symptoms back to baseline. Commonly reported post-acute COVID-19 symptoms are not specific to COVID-19 and are commonly reported regardless of infection status. It is not therefore expected that any group would report 0 incidence of such symptoms. However, the absence of difference in symptoms frequency among those who received two doses and those who were never infected suggests the excess reporting of these symptoms associated with COVID-19 infection in unvaccinated individuals is eliminated by vaccination. These associations were largely not seen among individuals who received a single dose of a COVID-19 vaccine, who were in most cases likely to have been infected prior to vaccination The results are consistent with the few other available studies on the topic reporting lower reporting of long-term symptoms following COVID-19 [17,19,21]. It also brings previously unavailable nuance on the effect of vaccination on reported long-term symtpoms, in terms of comparison to baseline incidence of such symtpoms, symptom specificity and time from acute infection. Approximately a third of participants in the current study continued to report post-COVID-19 symptoms, mostly fatigue, headache, and weakness. Our findings are consistent with a recently published study reporting similar proportions of post-COVID-19 symptoms [22]. An age-stratified analysis showed that the associations reported in the current study were largely confined to older age groups (above 60 years), a finding not consistent with a previous study reporting that the protective effect was confined to younger age groups [17]. Our analysis does not allow us to establish why the beneficial effect of the COVID-19 vaccine on long-term symptoms seems to be stronger in older age groups. A plausible explanation for the association found in the older individuals could be that younger individuals have more physiological reserve and are therefore able to recover on their own, which is not the case in older adults. Other studies have reported that frailty was associated with both age and worse outcomes following SARS-CoV-2 infection [23].

The unvaccinated and the vaccinated groups were comparable in sociodemographic characteristics except for age, reflecting vaccine coverage in the Israeli population [24]. As a result, some chronic conditions present at baseline were more common in the vaccinated group. However, since these conditions are more common in the older group who received two doses, we would anticipate even bigger differences between the vaccinated and unvaccinated group had we taken them into account in the analysis. This difference in age between the vaccinated and the unvaccinated individuals is consistent with the COVID-19 vaccination strategy implemented by the Israeli Ministry of Health, which targeted the older population during the initial stages of the COVID-19 vaccination rollout. The vaccinated and unvaccinated populations differed in terms of the proportion of asymptomatic individuals at acute COVID-19 presentation. A higher proportion of those who received two doses were asymptomatic at the time of diagnosis compared to the unvaccinated group, and those who were symptomatic at baseline reported less symptoms compared with those unvaccinated. These figures reflect the protection against symptomatic disease conferred by vaccines, which may also partly explain the lower proportion of reporting long-term symptoms among those vaccinated. However, the protective effect of vaccination against long-term symptoms persisted after adjusting for asymptomatic disease, suggesting a reduction in reported long COVID symptoms even among those symptomatic at the time of infection. The median follow-up duration was longer in the unvaccinated group than the vaccinated individuals. This is consistent with the fact that our follow-up period started 9 months before vaccines became available in Israel. One could therefore expect less symptoms in the unvaccinated as a result of having a longer time period to recover; however, the opposite was observed, and in any case the protective effect of vaccination remained after adjusting for follow-up time. The adjusted model suggested an even stronger protective effect of vaccination against reported long-term symptoms. However, we believe that the study was underpowered to detect age specific differences using an adjusted regression model.

Our study relied on self-reported positive PCR results. In Israel, most individuals have been PCR-tested multiple times for a range of reasons including being symptomatic, travelling, contact tracing, and screening. Specimens for the same patients are tested in different laboratories. Therefore, using positive lab results from a selection of laboratories (rather than all laboratories in the country) could lead to misclassification. Considering the consequences on daily life, including quarantine, it is unlikely that someone would forget having tested positive for SARS-CoV-2, or vice versa. It is also important to note that the vaccination policy in Israel, at the time of the survey, specified that SARS-CoV-2-infected individuals were in theory only eligible for a single dose of vaccine. Therefore, individuals who received one dose also differed from those who received two doses in terms of the sequence of events: while those who received two doses will have mostly been infected after having been vaccinated, many among those who received a single dose will have been infected prior to vaccination. Although our data does not allow to establish this sequence of events for each individual, the follow-up time and symptoms at presentation reflect this difference. Infection prior to vaccination could partially explain the observed lack of effect of one dose of vaccine regarding long-term reported symptoms. The effect of vaccination on long-term sequelae according to the infection/vaccination sequence warrants further research. With few patients reporting having been hospitalised, our cohort reflects the mild end of the COVID-19 spectrum, and the results cannot necessarily be extrapolated to patients who were more severely ill (and hospitalised) in the acute phase of the illness. Our study did not include children who are less likely to develop severe acute illness following infection but do report long-term effects [25]. Prevention of long-term symptoms could be one of the most important benefits of vaccination in this age group. At the time of this study, children were not eligible for COVID-19 vaccines in Israel. A final limitation is the self-reported nature of the symptoms, in particular since individuals who are eager to vaccinate may differ from those who do not vaccinate in terms of perceptions of health and illness.

This study offers a snapshot of the effectiveness of COVID-19 vaccine against the long-term effects of SARS-CoV-2 infection. The findings suggest that in addition to preventing severe disease and death [26], COVID-19 vaccines, or at least the BNT162b2 mRNA vaccine used in Israel, may play a critical role in preventing long-term outcomes of SARS-CoV-2 infection that fit within the current WHO definition of long COVID, in particular among older adults. Studies such as this one should be complemented by studies objectively measuring long-term health outcomes in COVID-19 patients in a clinical setting. Our cohort, which continues to recruit participants, will enable us to report with more precision whether the BNT162b2 mRNA protective effect reported in this study is sustained, as well as the effect of the different SARS-CoV-2 variants and vaccination on post-COVID symptoms.

## Data Availability

All data produced in the present study are available upon reasonable request to the authors

## Acknowledgements

We wish to thank Yehudit Hakmon, Eliran levi, Zion Levi, Nissim Neeman (Ziv Medical Centre), Mark Lifishitz, and Shelly Shalem (Galilee Medical Centre) for their technical help and support in disseminating the questionnaire.

---

**Figure 1.**
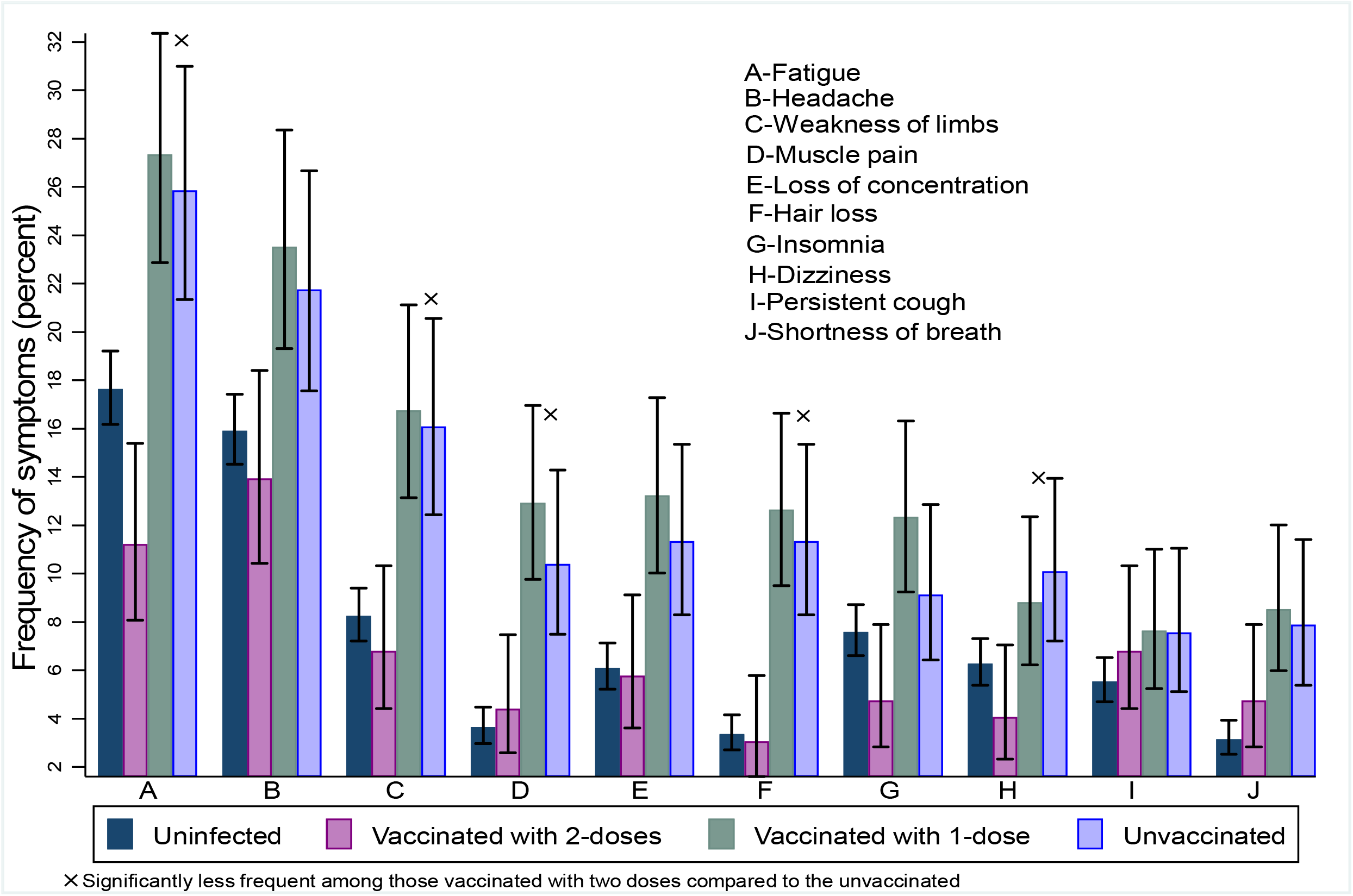
Frequency of most reported symptoms among the uninfected, the vaccinated and the unvaccinated

